# The association between family structure and adolescent physical activity levels: A systematic review of literature published since 2010

**DOI:** 10.1101/2023.07.04.23292220

**Authors:** Elena Mylona, Maartje Kletter, Helen M Jones, Marie Murphy, Richard Lampard, Oyinlola Oyebode

## Abstract

**Background:** Adolescent physical activity is influenced by biological, psychological, sociocultural, and environmental factors; however, no review has yet explored the effect of family structure (usually defined based on the relationships between people living in a household) on adolescent physical activity levels.

**Methods:** Databases MEDLINE, EMBASE, Web of Science, PsycINFO, CINAHL, and Sociological Abstracts were searched for peer-reviewed studies with a quantitative component published since 2010, with no restrictions on language, country, and year of data collection. Study screening, data extraction, and quality assessment occurred in duplicate. SWiM guidelines guided the narrative synthesis. PROSPERO protocol CRD42020221090.

**Results:** Thirty studies met inclusion criteria: 17 looked at global physical activity, 13 at leisure physical activity, and sport participation. All studies used cross-sectional designs and 27 assessed outcomes through a survey. Sixteen (10 of good quality) reported a significant association between family structure and adolescent physical activity. Of these, three did not specify the direction of this association while nine found adolescents in ‘traditional’ (two-parent) families were more physically active compared with other family structures. This association was stronger in studies of leisure-time physical activity. Two studies reported that adolescents with single mothers achieve more physical activity than adolescents living with neither parent. Two studies, focused on school physical exercise classes and active transport, found adolescents in single-parent households engaged in more physical activity than those living with two parents.

**Conclusion:** High-quality accelerometery, time diary, and longitudinal studies are needed to investigate the effect of family structure on adolescent physical activity and health sequelae. An improved understanding of social determinants of adolescent physical activity could inform health promotion strategies.

## BACKGROUND

Physical inactivity is a leading cause of several noncommunicable diseases like obesity, coronary heart disease and type 2 diabetes, increasing the risk of chronic illnesses and early deaths (1, 2). Promoting physical activity is a public health priority for the World Health Organisation (WHO), aiming at improving individuals’ physical and mental health, as well as creating more sustainable societies, as part of the Sustainable Development Agenda 2030 (3). Based on an international study funded by WHO that surveyed 1.6m students from 146 countries, adolescents aged 11-17 appear to be rather inactive, with four-fifths failing to meet recommended physical activity levels (4). Behavioural patterns and habits formed at a young age tend to carry over into adulthood, so encouraging physical activity for children and adolescents is paramount (5).

A systematic review of reviews shows that the amount of physical activity a person achieves is influenced by biological, psychological, sociocultural, and environmental factors (6). Published reviews have focused on several of these factors, their sub-factors, and their associations with physical activity levels (7, 8); however, no review has explored explicitly the effect of family structure on adolescent physical activity levels.

It is thought that children who grow up in a non-traditional/non-nuclear family structure, broadly defined as a non-two-parent biological/adoptive household, tend to experience more time and financial constraints, as well as less parental involvement and support; however, this has not been thoroughly investigated in terms of physical activity engagement (9, 10). With non-traditional, broadly referred to as ‘diverse’, family structures prevalent and proliferating (11), it is important to study the effect of family structure, as a major social determinant of various adolescent health outcomes.

A preliminary search using MEDLINE, PROSPERO and Cochrane Database of Systematic Reviews was conducted, to ensure that there was no current or in-progress review on this topic. This systematic review aims to identify and synthesize evidence on the effect of family structure on adolescent physical activity levels in quantitative observational cohort and cross-sectional studies, published since 2010, given the quality of life improvement of some forms of non-traditional families in the past decade due to changing (increasingly liberal) social attitudes (12). In the past, some of the barriers to physical activity for adolescents in non-traditional families may have been related to social conversative attitudes and these may no longer be relevant.

## METHODS

This systematic review has been registered with the International Prospective Register of Systematic Reviews (PROSPERO) (registration number: CRD42020221090) and was conducted according to the Preferred Reporting Items for Systematic Reviews and Meta-Analyses (PRISMA) and Synthesis without meta-analysis (SWiM) guidelines (13, 14).

### Search strategy

A search of MEDLINE using the key terms “adolescence”, “family structure” and “physical activity” was used to identify relevant articles. Key terms found in the titles, abstracts and index terms were used in development of the full MEDLINE strategy. Searches were developed with the advice of an academic librarian.

We searched six databases: MEDLINE, EMBASE, Web of Science, PsycINFO, CINAHL and Sociological Abstracts for studies published from 2010 up to and including the 8^th^ of February 2022, with no language restriction imposed. The search strategy was adapted for each of the included databases, including all identified key terms and the use of index terms where available. The detailed MEDLINE search strategy and terms can be found in the Additional file 1. Title and abstract screening was conducted independently in duplicate by EM, and MK, MM, HMJ and OO, using Rayyan.ai (15). Assessment of full-texts was performed independently in duplicate by EM, and MK and OO. Any discrepancies were resolved via discussion, and when necessary, through arbitration by OO.

### Eligibility criteria for inclusion

The population of interest is adolescents, defined by the WHO as individuals aged 10-19 (16). Studies using a wider age range were included only if the mean age was reported, or could be calculated from reported data, and fell in the 10-19 age range. This review included any peer-reviewed quantitative observational study, including prospective and retrospective cohort, and cross-sectional, studies. Mixed-method studies were also included if a quantitative component was present. Other study designs are included, if the studies look at the association of family structure and adolescent physical activity levels. Studies published in all languages were considered for inclusion.

Grey literature, case studies, reviews, editorials, PhD theses, conference abstracts, commentaries, and qualitative studies, were excluded.

### Exposure of interest

The exposure of interest is family structure. Family structure is defined as:

“A term that describes the members of a household who are linked by marriage or bloodline and is typically used in reference to at least one child residing in the home under the age of 18.” (17). This is often operationalised based on the number of parents (adoptive or related by blood) living in the household with the child.

### Outcomes

The outcomes of interest include any measure of physical activity behaviour, including, but not limited to, frequency (e.g. in days in the last or in a typical week), duration (e.g. in minutes/hours) or intensity (e.g. in METs (metabolic equivalents)). We also included studies examining sports participation, as a form as leisure time physical activity.

### Quality assessment

All studies retrieved were assessed independently by at least two reviewers (EM, MK, OO) for methodological quality using the National Institutes of Health (NIH) Study Quality Assessment Tool for Observational Cohort and Cross-Sectional Studies (18). This NIH tool uses 14 distinct criteria that are marked with “yes”, “no” or “cannot be determined” (not applicable or not reported). The results of the critical appraisal were reported by the reviewers who completed a quality evaluation table and rated each study as Good, Fair, or Poor based on their final score. Any disagreements between reviewers were resolved through discussion and involved a third reviewer, when necessary. Regardless of methodological quality, all studies meeting the inclusion criteria underwent data extraction and synthesis.

### Data synthesis

As anticipated, a meta-analysis was not conducted given the high heterogeneity of studies, mainly in terms of methods, age groups, family structure categories and physical activity outcomes. The Synthesis Without Meta-analysis (SWiM) guidelines were used to guide the narrative synthesis (14, 19).

### Changes in protocol

Despite not having a restricted year of publication in the protocol registration, the authors decided to look at studies published from 2010 onwards (without, however, excluding studies in which the data collection took place before 2010), given the quality of live improvement of some forms of non-traditional families in the past decade, as their rates of social exclusion, housing and material deprivation and low working intensity have decreased compared to previous years (12). The search strategy in Sociological Abstracts was applied only to titles, abstracts, and key-words, as applying the searches to full-texts returned a very large number of irrelevant studies because of the use of our search terms in other contexts within the discipline (e.g.: “exercise” of power). For similar reasons the database Scopus was not used despite being in the protocol registration.

## Results

### Study selection and characteristics

The literature search identified 15151 records and after removing the duplicates, 9361 underwent title and abstract screening allowing 9233 further studies to be excluded. In total, 128 full-text articles were assessed for eligibility and 30 met the inclusion criteria. The study selection process can be found in Figure 1.

**Figure 1.**
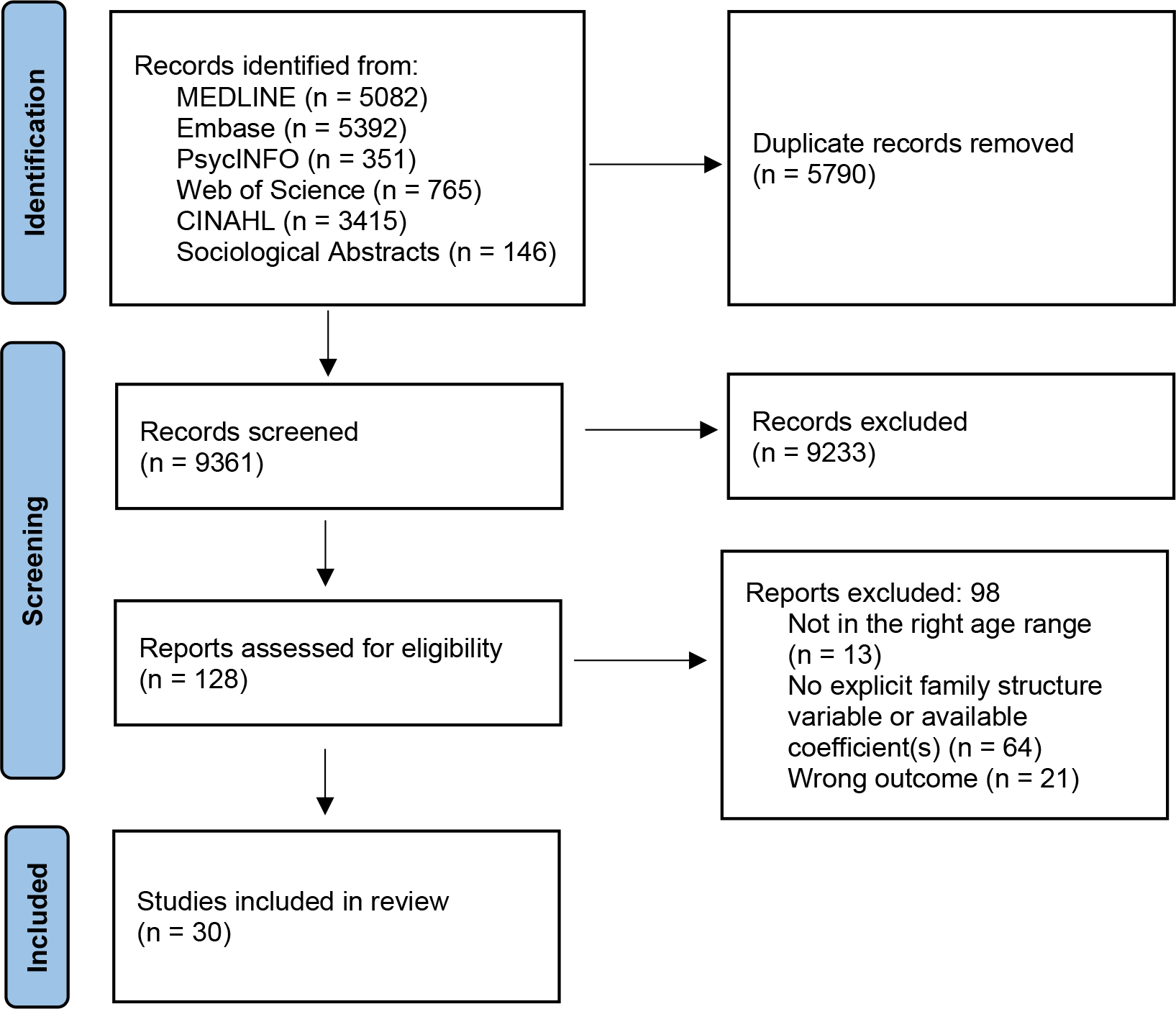
PRISMA flow chart.

Studies included in this systematic review examined family structure and its association with adolescent physical levels; however, the studies differed in their focus, with 17 looking at global physical activity (i.e.: physical activity accrued in leisure time as well as at school, and for travel) and 13 looking at leisure-time physical activity and/or sport participation only (Tables 1 and 2 respectively, show the studies’ characteristics).

**Table 1.**
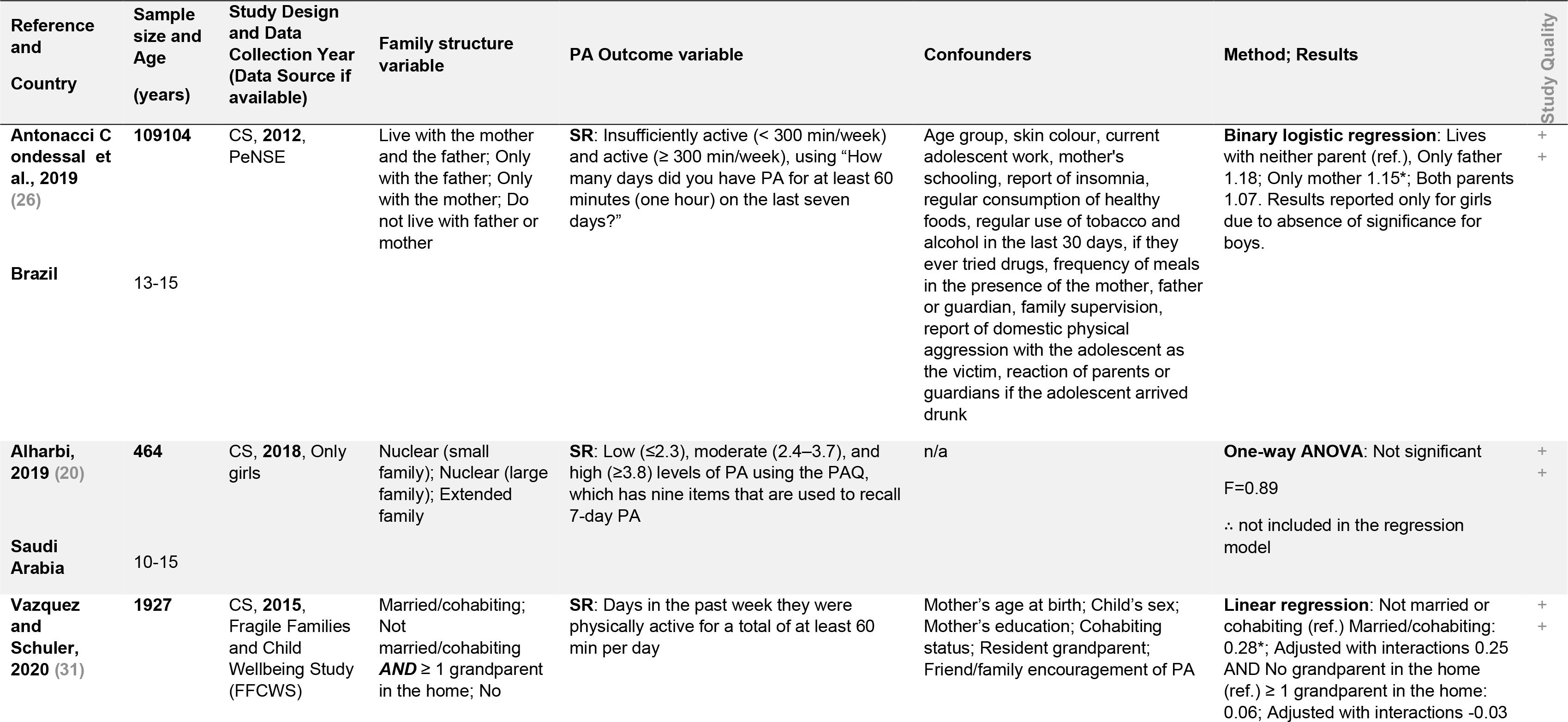
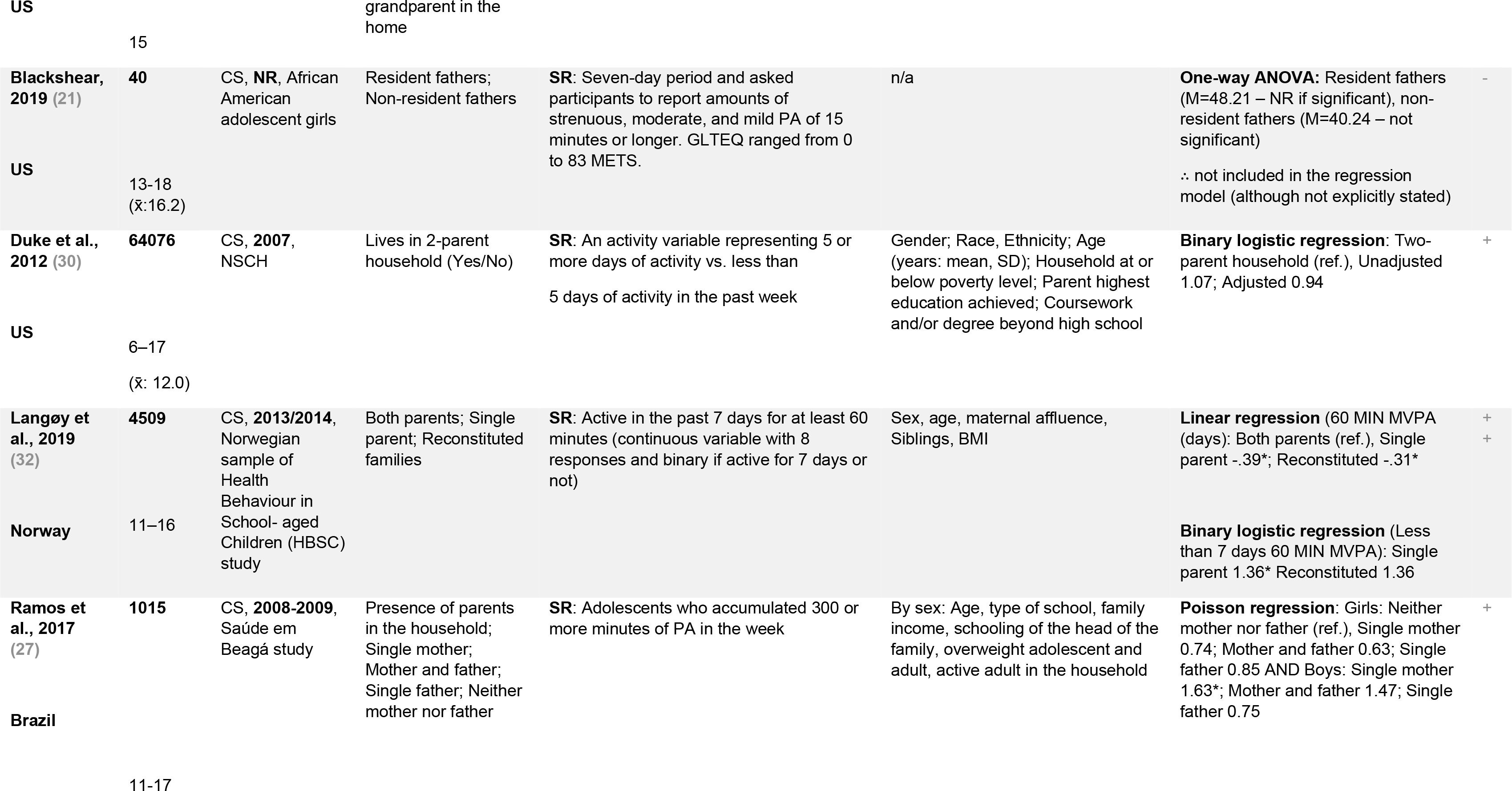
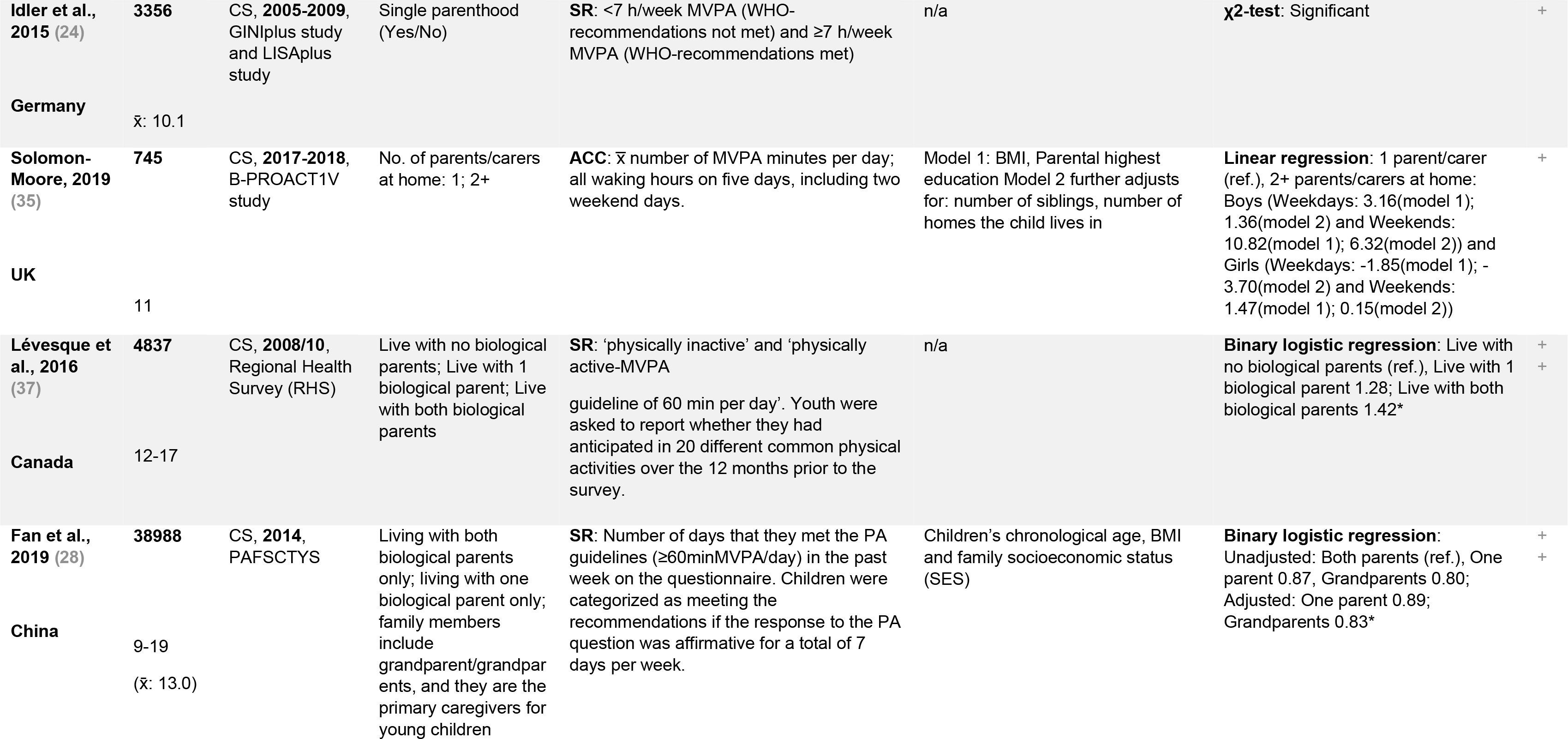
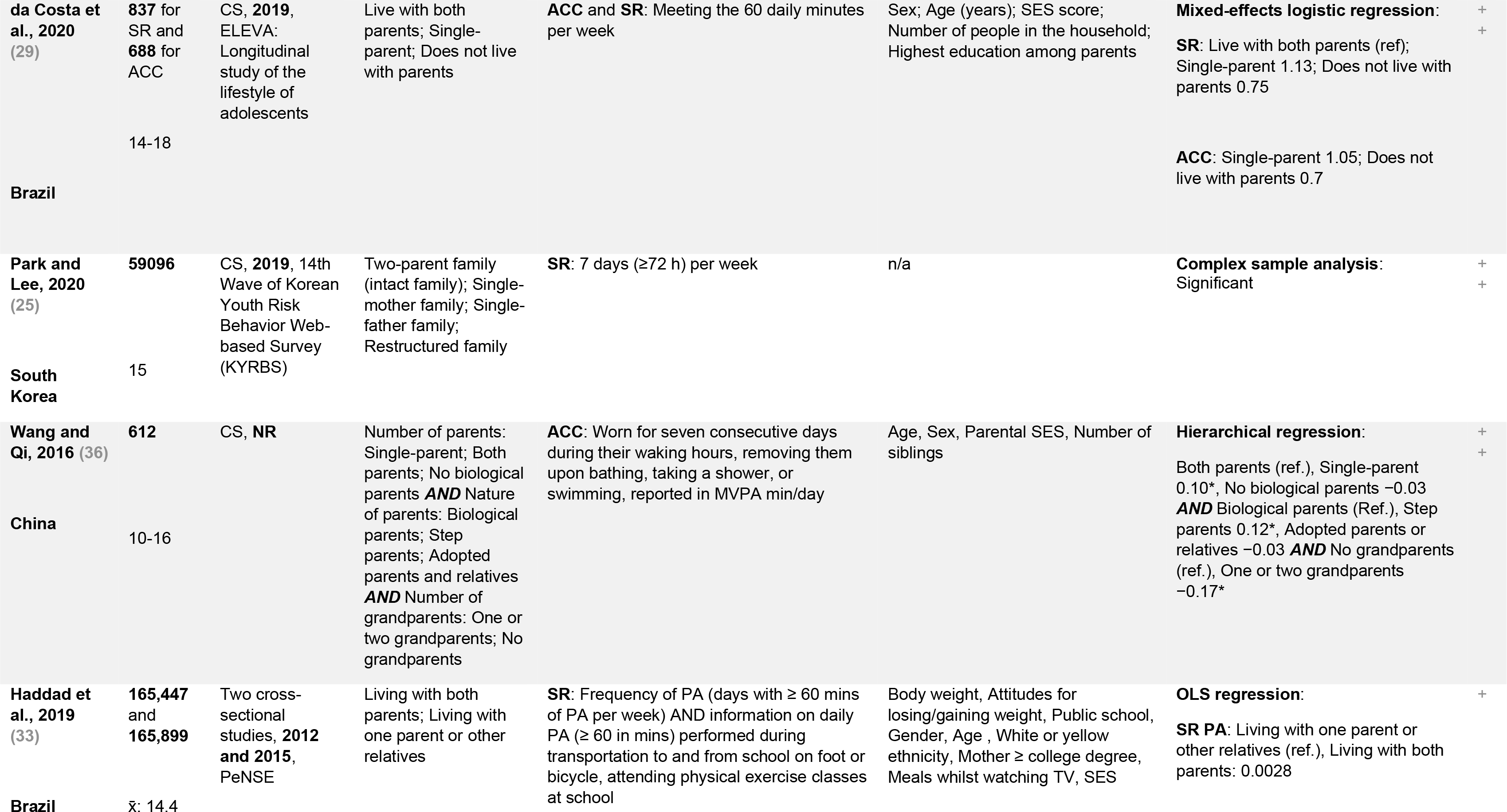
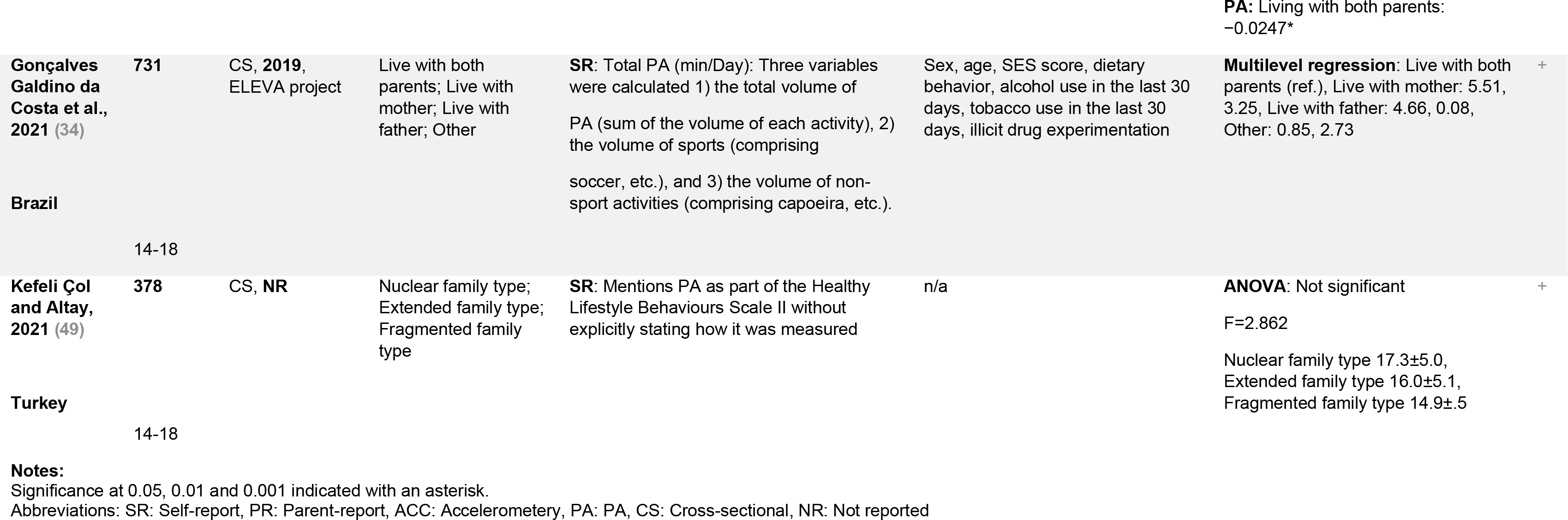
Global Physical Activity.

**Table 2.**
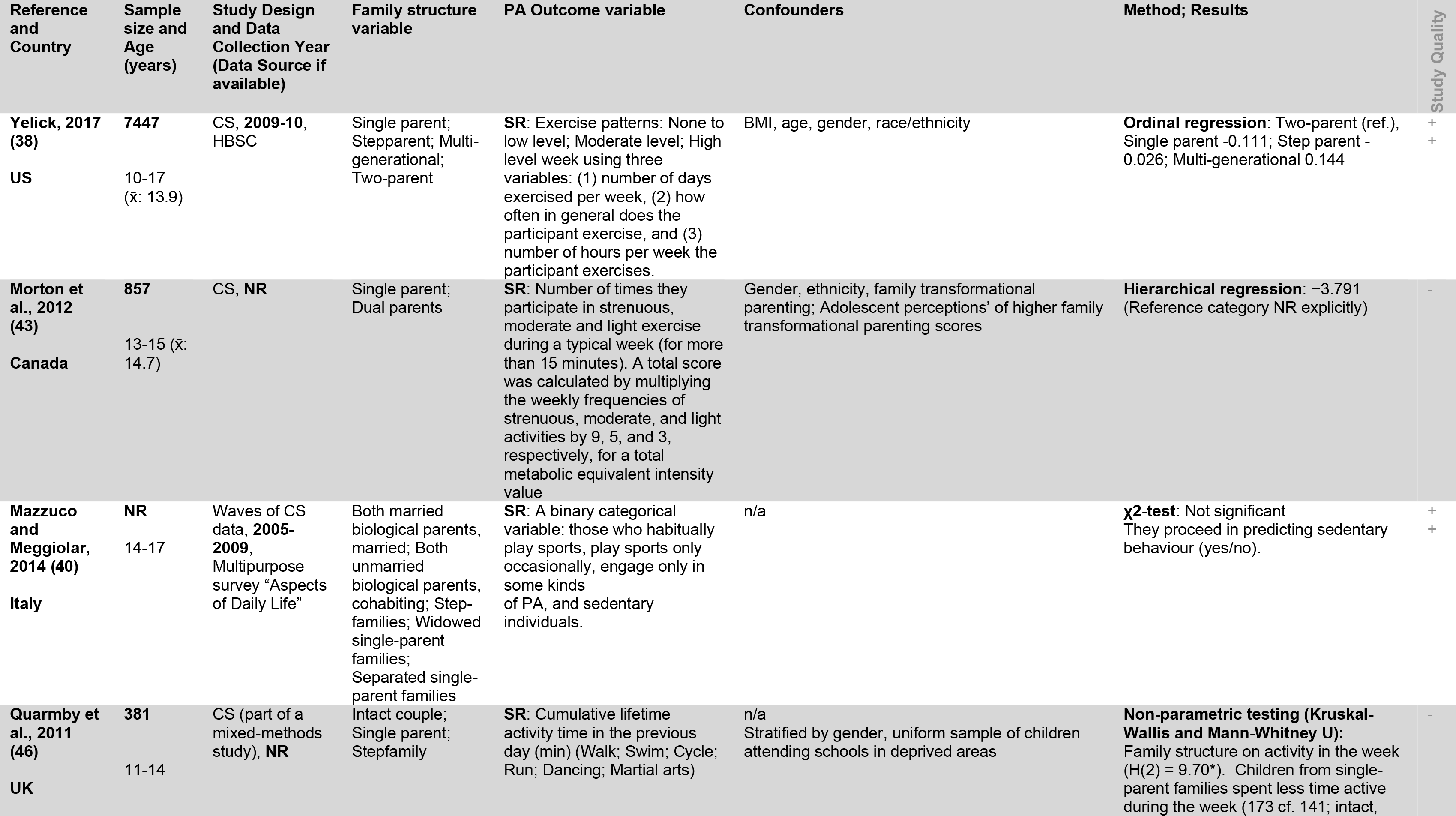
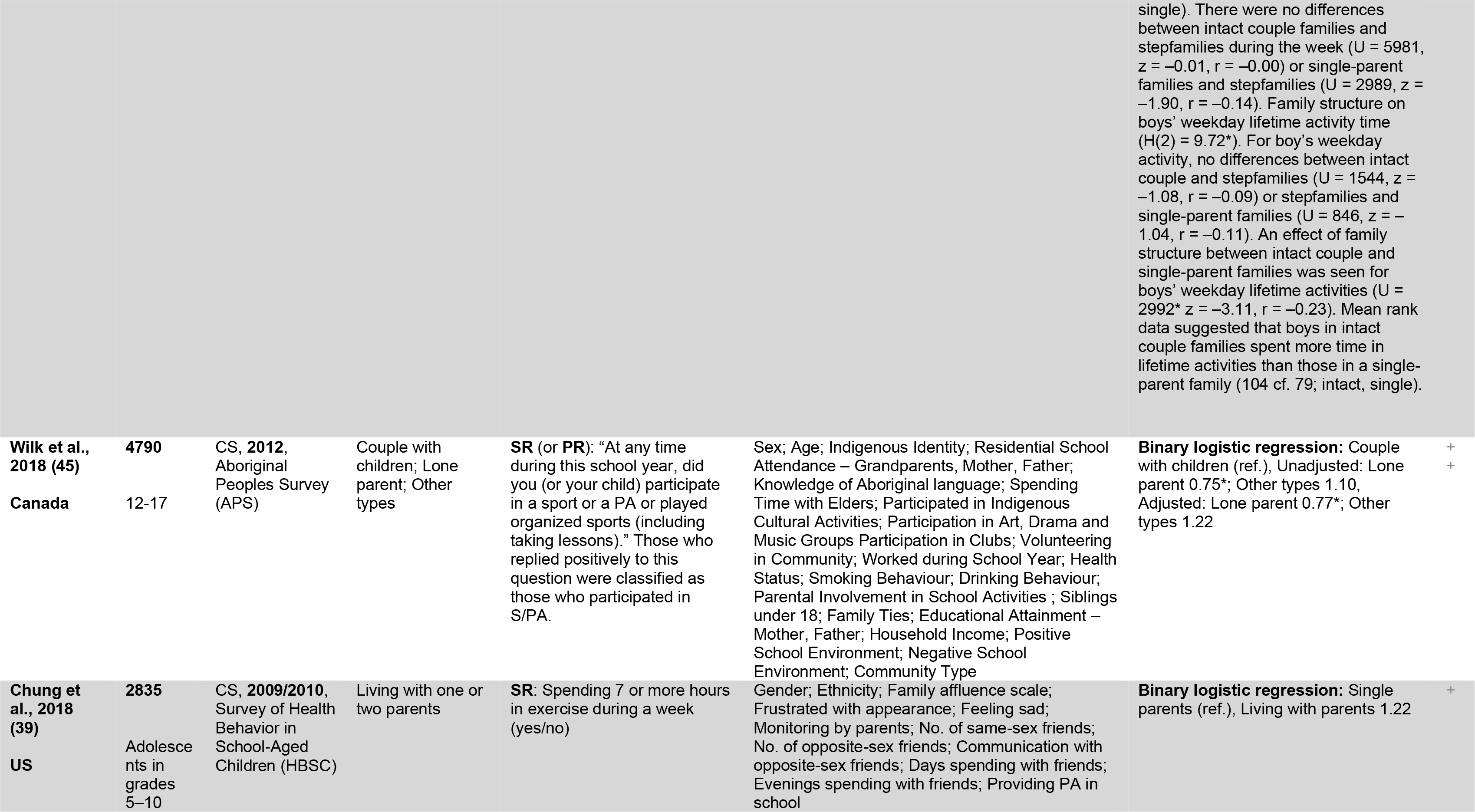
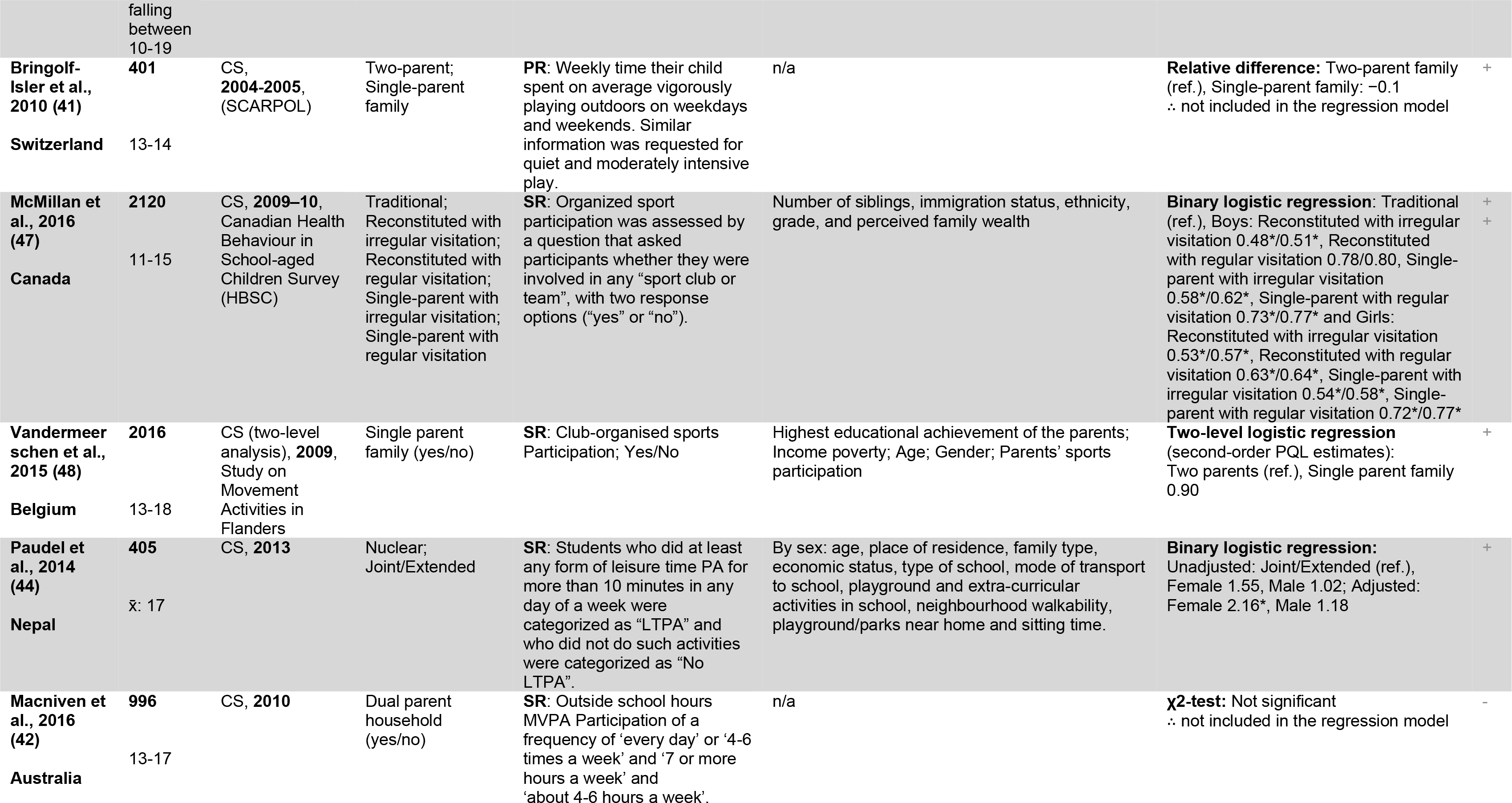
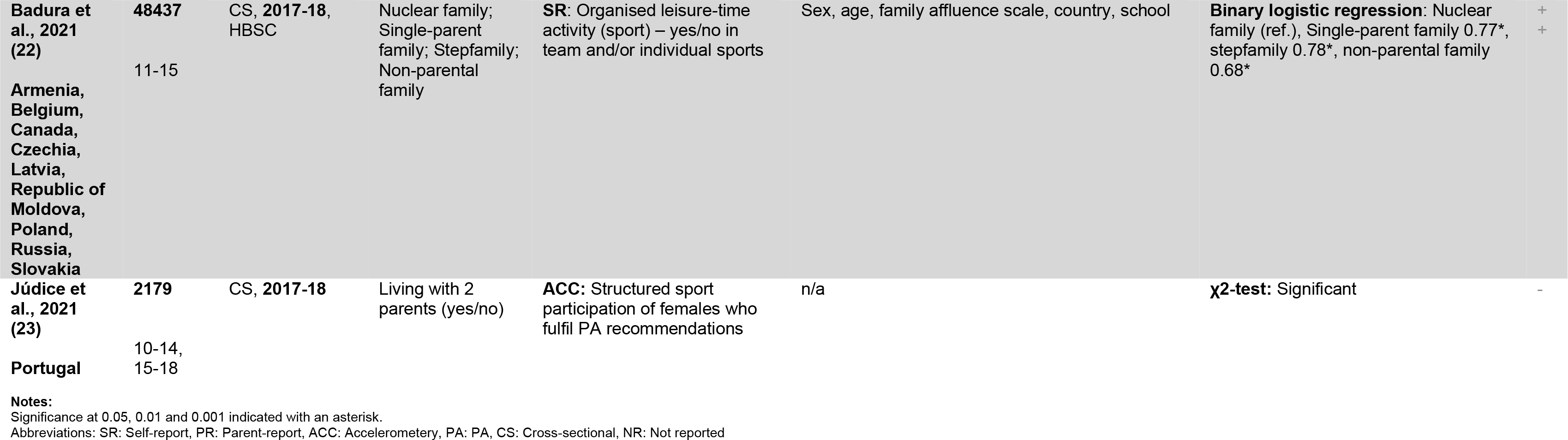
Leisure Physical Activity and Sport Participation.

For global physical activity, most studies were conducted in Brazil (n=5), followed by the United States (US) (n=3); China (n=2); and Saudi Arabia, Norway, Germany, United Kingdom (UK), Canada, South Korea, and Turkey (n=1). The sample sizes ranged from 40 to 64076, and most studies used secondary analysis of existing regional/national datasets (n= 13). There were two studies that only looked at females (20, 21).

For leisure physical activity and sport participation, most studies were conducted in Canada (n=3), followed by the US (n=2); and UK, Italy, Switzerland, Belgium, Nepal, and Australia (n=1). Only one study included populations from (nine) different countries (22) and one looked at females only (23). The sample sizes ranged from 381 to 48437 for single country studies, and 165899 for the cross-national study (22), and the majority of studies used secondary analysis of existing regional/national datasets (n= 8). Age ranges and means varied across studies.

### Risk of Bias

Risk of bias was assessed using the NIH Study Quality Assessment Tool for Observational Cohort and Cross-Sectional Studies (18), and 14 studies were classified as good, 11 as fair and five as poor. Half of the studies used a variable with more than two family structure categories, and 90% of studies used a validated instrument for measuring physical activity or accelerometery. Approximately one third of the studies controlled for all relevant key confounders: gender, a measure of socio-economic status, ethnicity, and age (or specified the population of interest the sample was drawn from). The individual quality score of each study is presented in Tables 1 and 2, and the detailed quality assessment table can be found in the Additional file 1.

## Global Physical Activity

### Meeting the weekly guidelines

Seven studies examined the association between family structure and meeting weekly guidelines, ≥ 60 minutes a day, either every day or 5 days a week, or ≥ 300 minutes a week. Five studies reported significant findings, with two studies, one (good) in South Korea (mean age 15) and one (fair) in Germany (mean age 10.1), reporting an association between family structure and meeting the weekly guidelines (24) (25). Two studies conducted in Brazil, with one (good) looking at 13-15 year olds (fully-adjusted model segregated by gender, findings were only significant for girls) (26) and one (fair) at 11-17 year olds (27), reported that adolescents living with their mothers only have higher odds of meeting the guidelines when compared to adolescents living with neither parent. A (good) study in China, looking at 9-19 year olds, reported that adolescents living with their grandparents as their primary caregivers have lower odds of meeting the weekly guidelines (28).

Two studies reported no statistically significant findings, one (good) study from Brazil using accelerometery and self-report for 14-18 year olds (29), and a (fair) US study (mean age 12) (30).

### Days of ≥60 daily minutes of moderate-to-vigorous physical activity (MVPA) in a week

Two good quality studies, one conducted in the US (age 15) (31), and one in Norway (11-16 years) (32), used multivariable regression to predict the amount of physically active days (≥60 daily minutes) per week. Both studies reported significant associations between family structure and this outcome, showing that adolescents living with both parents are more likely to be physically active for more days a week (compared to living with not married/cohabiting parents, and living with a single parent and reconstituted families respectively). A fair quality study conducted in Brazil (mean age 14.4) reported no statistically significant findings (33).

### Daily minutes of MVPA

Four studies looked specifically at daily minutes of MVPA. Three reported no significant findings; a (poor) US study of African American girls aged 13-18, exploring the association with having a resident father or not (21), a (fair) Brazilian study looking at the total daily minutes of physical activity of 14-18 year olds (34), and a (fair) UK study using accelerometery to predict daily MVPA minutes at age 11 (35). In contrast, a (good) accelerometery study conducted in China (36) using multivariable regression, showed that 10-16 year old adolescents in single parent families are more likely to engage in more MVPA daily minutes per week, when compared to those living with both parents; the same holding true for those living with stepparents, when compared with those living with their biological parents, and for those who live with no grandparents, when compared with those living with one or two grandparents.

### Yearly MVPA

A (good) Canadian study examined 12-17 year olds achieving 60 daily minutes of MVPA over the last 12 months. The study only showed an unadjusted model for family structure, reporting that adolescents living with both parents have higher odds of achieving 60 daily minutes of MVPA in a year, when compared to those living with no biological parents (37).

### Low, moderate, or high levels of Physical Activity

A (good) study looked at the association between family structure and 10-15 year old girls’ engagement in low, moderate, or high levels of physical activity in Saudi Arabia, reporting no statistically significant results (20).

## Leisure Physical Activity and Sport Participation

### Leisure Physical Activity

In total, nine studies looked at leisure physical activity. Six studies reported no significant findings: two were conducted in the US, one of good (38) and one of fair quality (39), looking at 10-17 and 10-19 year olds respectively, one (good) study from Italy (40), looking at 14-17 year olds, one (fair) in Switzerland (41), looking at 13-14 year olds, using parent-report, and two poor quality studies, one in Australia (42), looking at 13-17 year olds, and one in Canada (43), looking at 13-15 year olds.

A (fair) study conducted in Nepal, looked at 17-year-olds and ‘any form of leisure time PA for more than 10 minutes in any day of a week’. Statistically significant results were reported for females, showing them more likely to engage in more exercise when in nuclear families, compared with joint/extended families (44). A (good) Canadian study used a binary outcome for yearly involvement in leisure physical activity, and reporting that 12-17 year olds in traditional families are more likely to engage in more leisure time physical activity in a year compared with children in lone parent or other types of families (45). A (poor) study conducted in the UK looked at daily minutes spent on the previous day in leisure-time physical activity of 11-14 year olds. They report no significant findings for boys or girls on weekends, nor for girls on weekdays, but report that boys in intact couple families spend more time in leisure-time activities than those in a single-parent family (46).

### Organised Sport Participation

Four studies looked explicitly at organised sport participation (22, 23, 47, 48), three of which found a significant association between family structure and organised sport participation. A (good) study in Canada (47), a (fair) one in Belgium (48) and a (good) study using data from 9 different countries (22) used multivariable regression, with the last two reporting significant findings, showing that adolescents aged 11-15 in nuclear families have higher odds of participating in organised sports, when compared with other family structures, while the Canadian study reported no significant findings. A (poor) Portuguese study reported a significant association between family structure and organised sport participation of 10-14 and 15-18 year old females who fulfil physical activity recommendations, using accelerometery (23).

## Other outcomes

### Active travelling to school and Physical Education

A (fair) study conducted in Brazil (33) reported significant findings, with adolescents (mean age 14.4) living with one parent or other relatives doing slightly better compared to those living with both parents when it comes to daily physical activity (≥ 60 in minutes) performed during transportation to and from school on foot or bicycle and/or attending physical education classes at school.

### Unspecified outcome

A (poor) study conducted in Turkey included data on physical activity collected as part of the ‘Healthy Lifestyle Behaviours Scale II’, without explicitly stating how it was measured, reporting no significant association between family structure and physical activity of 14-18-year-olds (49).

## Discussion

This review has examined evidence on the association between family structure and adolescent physical activity levels. Despite ample research on social determinants of adolescent physical activity levels, only 30 quantitative studies published since 2010 included family structure as one the determinants examined. Evidence was gathered from cross-sectional observational studies, from a range of different countries.

There is some evidence (16 studies, 10 of good quality) of an association between adolescent physical activity and family structure. Of these 16 studies, three of mixed quality did not specify the effect size or direction of this association (23–25). Nine (seven of which good quality), showed that adolescents in ‘traditional’ families engage in more physical activity when compared to other family structures (22, 28, 31, 32, 37, 44–47). This association was stronger in studies of leisure time physical activity. In addition, two studies (one good, one fair) reported that adolescents with single mothers are more likely to report higher levels of physical activity, when compared to adolescents living with neither parent (26, 27). However, one fair study found adolescents living in single-parent households engaged in more physical exercise classes at school (33), and a further good study found that adolescents in single-parent households undertook more active travel to and from school, when compared with those living with two parents (36).

These findings suggest that adolescents, ages ∼14-17, growing up in single-parent households in some contexts may undertake less physical activity than adolescents in traditional family structures. However, it is possible that the association between family structure and physical activity varies depending on the type of activity, with those in non-traditional families being less likely to undertake leisure time physical activity and more likely to accrue active travel time (9, 10). This might be due to time pressures of financial pressures for a single parent making it harder to support an adolescent to access or afford leisure time physical activity opportunities.

Most studies used self-report questionnaires, and even though most used validated instruments, they are prone to measurement error. Therefore, many of these studies were identified as ‘good’, despite inherent weaknesses due to their study designs. There is thus an evidence gap that requires employing measures less prone to measurement error to estimate the effect of family structure on adolescent physical activity, such as accelerometery studies or studies using time-diary data, but also studies using longitudinal data, to provide stronger evidence to support future policy decisions.

A major strength of this systematic review is the rigorous approach to identifying literature, including consulting an academic librarian to develop the search strategy. Selection of studies and quality appraisal were conducted in duplicate by two independent reviewers, with data extraction conducted by one reviewer and checked by a second, improving the reliability of the data synthesis. This review has possibly missed evidence published in grey literature, as it focused on studies in peer-reviewed journals, published since 2010, aiming at synthesizing the latest available evidence to reflect social changes and adolescent activity patterns changing across decades. Included studies were also restricted to quantitative analysis, potentially missing out on qualitative evidence. However, this systematic review is to date the most comprehensive, with searches conducted in six different databases, covering a wide range of Social Science literature, not only the field of Health Science.

### Conclusion

Based on the available evidence, it is unclear whether adolescents in traditional family structures do better in terms of physical activity, when compared with adolescents living in (broadly defined) ‘non-traditional’ family structures. There is evidence that adolescents in nuclear families engage in more leisure time physical activity. Those designing or implementing interventions to increase leisure time physical activity should consider how feasible or accessible these interventions are to adolescents from non-traditional family structures. It is possible that any causal effect of family structure on adolescent physical activity levels is specific to the wider social and cultural context which would explain the lack of consistency in the literature identified. More studies employing measures less prone to measurement error are required to estimate the effect of family structure on adolescent physical activity, such as more accelerometery studies or studies using time-diary data. There is also a substantial evidence gap when it comes to long-term effects, requiring stronger evidence to support future policy decisions. The need for this evidence is likely to be growing, as the number of non-traditional families continues to increase.

## Declarations

### Ethical approval

No ethical approval was required for this study as data used were obtained from previously peer-reviewed published studies.

## Authors’ contributions

EM: Study concept and design, literature searches, full-text screening, data extraction, quality ratings, interpretation of data, drafting and writing of manuscript, critical revision of manuscript. MK: Study screening, full-text screening, quality ratings. HJ: Study screening and critical revision of manuscript. MM: Study screening and critical revision of manuscript. RL: Interpretation of data and critical revision of manuscript. OO: Study concept and design, study screening, quality ratings, interpretation of data and critical revision of manuscript. All authors have reviewed the final manuscript submitted for publication.

## Supporting information

Additional File 1. MEDLINE Search Strategy and Quality Appraisal

## Data Availability

Data used were obtained from previously peer-reviewed published studies.

## Acknowledgements

We would like to thank Ms Samantha Johnson, our academic librarian, who has helped in developing the search strategy for this study’s protocol.

## References

1. Ding D, Lawson KD, Kolbe-Alexander TL, Finkelstein EA, Katzmarzyk PT, van Mechelen W, et al. The economic burden of physical inactivity: a global analysis of major non-communicable diseases. The Lancet. 2016;388(10051):1311–24.

2. Stamatakis E, Ding D, Ekelund U, Bauman AE. Sliding down the risk factor rankings: reasons for and consequences of the dramatic downgrading of physical activity in the Global Burden of Disease 2019. British Journal of Sports Medicine. 2021;55(21):1222.

3. WHO. Global action plan on physical activity 2018–2030: more active people for a healthier world 2018 [Available from: https://www.who.int/publications/i/item/9789241514187.

4. Guthold R, Stevens GA, Riley LM, Bull FC. Global trends in insufficient physical activity among adolescents: a pooled analysis of 298 population-based surveys with 1.6 million participants. The Lancet Child & Adolescent Health. 2020;4(1):23–35.

5. Frech A. Healthy Behavior Trajectories between Adolescence and Young Adulthood. Adv Life Course Res. 2012;17(2):59–68.

6. Sterdt E, Liersch S, Walter U. Correlates of physical activity of children and adolescents: A systematic review of reviews. Health Education Journal. 2013;73(1):72–89.

7. Craggs C, Corder K, van Sluijs EM, Griffin SJ. Determinants of change in physical activity in children and adolescents: a systematic review. Am J Prev Med. 2011;40(6):645–58.

8. Fisher A, Smith L, van Jaarsveld CH, Sawyer A, Wardle J. Are children’s activity levels determined by their genes or environment? A systematic review of twin studies. Prev Med Rep. 2015;2:548–53.

9. Fallesen P, Gähler M. Family type and parents’ time with children: Longitudinal evidence for Denmark. Acta Sociologica. 2019;63(4):361–80.

10. Culliney M, Haux T, McKay S. Family structure and poverty in the UK: An evidence and policy review. University of Lincoln; 2014.

11. Pearce LD, Hayward GM, Chassin L, Curran PJ. The Increasing Diversity and Complexity of Family Structures for Adolescents. J Res Adolesc. 2018;28(3):591–608.

12. Policy Department for Citizens’ Rights and Constitutional Affairs. The situation of single parents in the EU. 2020.

13. Moher D, Liberati A, Tetzlaff J, Altman DG. Preferred reporting items for systematic reviews and meta-analyses: the PRISMA statement. BMJ. 2009;339:b2535.

14. Campbell M, McKenzie JE, Sowden A, Katikireddi SV, Brennan SE, Ellis S, et al. Synthesis without meta-analysis (SWiM) in systematic reviews: reporting guideline. BMJ. 2020;368:l6890.

15. Ouzzani M, Hammady H, Fedorowicz Z, Elmagarmid A. Rayyan—a web and mobile app for systematic reviews. Systematic Reviews. 2016;5(1):210.

16. WHO. Adolescent health [Available from: https://www.who.int/health-topics/adolescent-health#tab=tab_1.

17. Pasley K, Petren R, E. Family Structure: Encyclopedia of family studies, Wiley Online Library; 2015 [Available from: https://onlinelibrary.wiley.com/doi/10.1002/9781119085621.wbefs016.

18. NIH. 2021 [Available from: https://www.nhlbi.nih.gov/health-topics/study-quality-assessment-tools.

19. Campbell M, Katikireddi SV, Sowden A, Thomson H. Lack of transparency in reporting narrative synthesis of quantitative data: a methodological assessment of systematic reviews. J Clin Epidemiol. 2019;105:1–9.

20. Alharbi M. Influence of individual and family factors on physical activity among Saudi girls: a cross-sectional study. Ann Saudi Med. 2019;39(1):13–21.

21. Blackshear T. Fathers – An Untapped Resource for Increasing Physical Activity among African American Girls. Montenegrin Journal of Sports Science & Medicine. 2019;8.

22. Badura P, Hamrik Z, Dierckens M, Gobina I, Malinowska-Cieślik M, Furstova J, et al. After the bell: adolescents’ organised leisure-time activities and well-being in the context of social and socioeconomic inequalities. Journal of Epidemiology and Community Health. 2021;75:jech-2020.

23. Júdice PB, Magalhães JP, Rosa GB, Henriques-Neto D, Hetherington-Rauth M, Sardinha LB. Sensor-based physical activity, sedentary time, and reported cell phone screen time: A hierarchy of correlates in youth. Journal of Sport and Health Science. 2021;10(1):55–64.

24. Idler N, Teuner CM, Hunger M, Holle R, Ortlieb S, Schulz H, et al. The association between physical activity and healthcare costs in children--results from the GINIplus and LISAplus cohort studies. BMC public health. 2015;15:437-.

25. Park H, Lee K-S. The association of family structure with health behavior, mental health, and perceived academic achievement among adolescents: a 2018 Korean nationally representative survey. BMC Public Health. 2020;20(1):510.

26. Antonacci Condessa L, Cardoso Chaves O, Marcelina Silva F, Carvalho Malta D, Teixeira Caiaffa W. Sociocultural factors related to the physical activity in boys and girls: PeNSE 2012. Revista de Saude Publica. 2019;53(1).

27. Ramos CGC, Andrade RG, Andrade ACS, Fernandes AP, Costa D, Xavier CC, et al. Family context and the physical activity of adolescents: comparing differences. Rev Bras Epidemiol. 2017;20(3):537–48.

28. Fan X, Zhu Z, Zhuang J, Liu Y, Tang Y, Chen P, et al. Gender and age differences in the association between living arrangement and physical activity levels among youth aged 9-19 years in Shanghai, China: a cross-sectional questionnaire study. BMC Public Health. 2019;19(1):1030.

29. da Costa BGG, Chaput J-P, Lopes MVV, Malheiros LEA, Tremblay MS, Silva KS. Prevalence and sociodemographic factors associated with meeting the 24-hour movement guidelines in a sample of Brazilian adolescents. PLOS ONE. 2020;15(9):e0239833.

30. Duke NN, Borowsky IW, Pettingell SL. Parent perceptions of neighborhood: relationships with US youth physical activity and weight status. Matern Child Health J. 2012;16(1):149–57.

31. Vazquez C, Schuler B. Adolescent Physical Activity Disparities by Parent Nativity Status: the Role of Social Support, Family Structure, and Economic Hardship. J Racial Ethn Health Disparities. 2020;7(6):1079–89.

32. Langøy A, Smith ORF, Wold B, Samdal O, Haug EM. Associations between family structure and young people’s physical activity and screen time behaviors. BMC Public Health. 2019;19(1):433.

33. Haddad MR, Sarti FM, Nishijima M. Association between selected individual and environmental characteristics in relation to health behavior of Brazilian adolescents. Eat Weight Disord. 2021;26(1):331–43.

34. da Costa BG, Chaput J-P, Lopes MV, Malheiros LE, Silva KS. Associations between Sociodemographic, Dietary, and Substance Use Factors with Self-Reported 24-Hour Movement Behaviors in a Sample of Brazilian Adolescents. International Journal of Environmental Research and Public Health. 2021;18(5).

35. Solomon-Moore E, Salway R, Emm-Collison LG, Sebire SJ, Thompson JL, Jago R. A Longitudinal Study of the Associations of Family Structure with Physical Activity across the Week in Boys and Girls. Int J Environ Res Public Health. 2019;16(20).

36. Wang L, Qi J. Association between Family Structure and Physical Activity of Chinese Adolescents. Biomed Res Int. 2016;2016:4278682.

37. Lévesque L, Janssen I, Xu F. Correlates of physical activity in First Nations youth residing in First Nations and northern communities in Canada. Can J Public Health. 2015;106(2):e29–e35.

38. Yelick A. The Effects of Family Structure on Consumption and Exercise Patterns for Adolescent Youth: C & A. Child & Adolescent Social Work Journal. 2017;34(4):381–95.

39. Chung SJ, Ersig AL, McCarthy AM. Parent, school, and peer factors related to U.S. adolescents’ diet and exercise. Journal for Specialists in Pediatric Nursing. 2018;23(4):e12227.

40. Mazzuco S, Meggiolaro S. Family Structures and Health Behaviour in Adolescents. Child Indicators Research. 2014;7(1):155–75.

41. Bringolf-Isler B, Grize L, Mäder U, Ruch N, Sennhauser FH, Braun-Fahrländer C. Built environment, parents’ perception, and children’s vigorous outdoor play. Prev Med. 2010;50(5- 6):251–6.

42. Macniven R, Hearn S, Grunseit A, Richards J, Nutbeam D, Bauman A. Correlates of physical activity among Australian Indigenous and non-Indigenous adolescents. Aust N Z J Public Health. 2017;41(2):187–92.

43. Morton KL, Wilson AH, Perlmutter LS, Beauchamp MR. Family leadership styles and adolescent dietary and physical activity behaviors: a cross-sectional study. International Journal of Behavioral Nutrition and Physical Activity. 2012;9(1):48.

44. Paudel S, Subedi N, Bhandari R, Bastola R, Niroula R, Poudyal AK. Estimation of leisure time physical activity and sedentary behaviour among school adolescents in Nepal. BMC Public Health. 2014;14(1):637.

45. Wilk P, Maltby A, Cooke M, Forsyth J. Correlates of Participation in Sports and Physical Activities among Indigenous Youth. Aboriginal Policy Studies. 2018;7(1).

46. Quarmby T, Dagkas S, Bridge M. Associations between children’s physical activities, sedentary behaviours and family structure: a sequential mixed methods approach. Health Educ Res. 2011;26(1):63–76.

47. McMillan R, McIsaac M, Janssen I. Family Structure as a Correlate of Organized Sport Participation among Youth. PLOS ONE. 2016;11(2):e0147403.

48. Vandermeerschen H, Vos S, Scheerder J. Who’s joining the club? Participation of socially vulnerable children and adolescents in club-organised sports. Sport, Education and Society. 2015;20(8):941–58.

49. Kefelİ ÇOl B, Altay B. The Effect of Family Characteristics of the Adolescents Upon Their Healthy Lifestyle Behaviors. Turkiye Klinikleri Journal of Nursing Sciences. 2021;13(3):564–71.

