## Additional File 1. MEDLINE Search Strategy and Quality Appraisal for "The association between family structure and adolescent physical activity levels: A systematic review of literature published since 2010"

**Table 1**. MEDLINE Search Strategy

| Concept | Free-text terms | MeSH terms |
|---|---|---|
| **Adolescents** | (adolescen*) OR (teen*) OR (young people) OR (young person*) OR (young adult*) OR (youth or youths) OR (student or students) OR (juvenile*) OR (underage) OR (girl or girls) OR (boy or boys) | Adolescent |
| **Physical activity** | (physical activ*) OR (physical exercise*) OR (MVPA) OR (activ*) OR (exercise*) OR (fitness) OR (muscular strength) OR (muscular endurance) or (explosive strength) OR (sport*) OR (athletic*) | Exercise<br>Sports |
| **Family structure** | (family structure*) OR (family type*) OR (single parent* or single-parent*) OR (lone paren* or lone-parent*) OR (lone mother* or lone-mother*) OR (lone father* or lone-father*) OR (two parent or two-parent) OR (step parent* or step-parent*) OR (step famil* or step-famil*) OR (cohabiting famil*) OR (adoptive famil*) OR (grandparent famil*) OR (foster care famil* or foster-care famil*) OR (blended famil*) OR (cohabiting parent*) OR (same sex famil* or same-sex famil*) OR (multi-generational famil*) OR (binuclear famil*) OR (*engaged famil*) OR (enmeshed famil*) OR (under organized famil* or under-organized famil*) OR (family stabilit*) OR (nuclear famil*) OR (non-nuclear famil*) OR (nonnuclear famil*) OR (biological parent*) OR (biological famil*) OR (adoptive parent*) OR (single parent*) OR (foster parent*) OR (step-famil* OR stepfamily*) OR (traditional famil*) OR (non-traditional family*) OR (nontraditional family*) | Nuclear family<br>Single parent |

**Table 2.** Quality appraisal of studies

| Reference | Study objective[1] | Study population[2] | 50% non-response rate[3] | Participants selected at the same time[4] | Sample size justification[5] | Exposure measured prior to outcome[6] | Sufficient timeframe for association[7] | More than two family structure categories[8] | Valid, reliable and consistent independent variable[9] | Exposure assessed more than once over time[10] | Valid, reliable and consistent outcome[11] | Blinded outcome[12] | 20% follow-up[13] | Key confounders (Sex, SES; age/ethnicity is appropriate)[14] | Quality Rating[15] |
|---|---|---|---|---|---|---|---|---|---|---|---|---|---|---|---|
| **Antonacci Condessal et al., 2019 [1]** | Yes | Yes | NR | Yes | No | No | No | Yes | Yes | No | Yes | NA | NA | Yes | Good |
| **Yelick, 2017 [2]** | Yes | Yes | Yes | Yes | No | No | No | Yes | Yes | No | Yes | NA | NA | No | Good |
| **Alharbi, 2019 [3]** | Yes | Yes | Yes | Yes | No | No | No | Yes | Yes | No | Yes | NA | NA | Yes | Good |
| **Vazquez and** | Yes | Yes | Yes | Yes | No | No | No | No | Yes | No | Yes | NA | NA | Yes | Good |

[1] Was the research question or objective in this paper clearly stated?
[2] Was the study population clearly specified and defined?
[3] Was the participation rate of eligible persons at least 50%?
[4] Were all the subjects selected or recruited from the same or similar populations (including the same time period)? Were inclusion and exclusion criteria for being in the study prespecified and applied uniformly to all participants?
[5] Was a sample size justification, power description, or variance and effect estimates provided?
[6] For the analyses in this paper, were the exposure(s) of interest measured prior to the outcome(s) being measured?
[7] Was the timeframe sufficient so that one could reasonably expect to see an association between exposure and outcome if it existed?
[8] For exposures that can vary in amount or level, did the study examine different levels of the exposure as related to the outcome (e.g., categories of exposure, or exposure measured as continuous variable)?
[9] Were the exposure measures (independent variables) clearly defined, valid, reliable, and implemented consistently across all study participants?
[10] Was the exposure(s) assessed more than once over time?
[11] Were the outcome measures (dependent variables) clearly defined, valid, reliable, and implemented consistently across all study participants?
[12] Were the outcome assessors blinded to the exposure status of participants?
[13] Was loss to follow-up after baseline 20% or less?
[14] Were key potential confounding variables measured and adjusted statistically for their impact on the relationship between exposure(s) and outcome(s)?

| | | | | | | | | | | | | | | |
|---|---|---|---|---|---|---|---|---|---|---|---|---|---|---|
| **Schuler, 2020 [4]** | | | | | | | | | | | | | | |
| **Morton et al., 2012 [5]** | Yes | Yes | NR | Yes | No | No | No | No | Yes | No | Yes | NA | NA | No | Poor |
| **Blackshear, 2019 [6]** | Yes | Yes | NR | Yes | No | No | No | No | Yes | No | Yes | NA | NA | No | Poor |
| **Duke et al., 2012 [7]** | Yes | Yes | NR | Yes | No | No | No | No | Yes | No | Yes | NA | NA | Yes | Fair |
| **Mazzuco and Meggiolar, 2014 [8]** | Yes | Yes | NR | Yes | Yes | No | No | Yes | Yes | No | Yes | NA | NA | No | Good |
| **Quarmby et al., 2011 [9]** | Yes | Yes | NR | Yes | No | No | No | Yes | Yes | No | No | NA | NA | No | Poor |
| **Langøy et al., 2019 [10]** | Yes | Yes | Yes | Yes | No | No | No | Yes | Yes | No | Yes | NA | NA | No | Good |
| **Wilk et al., 2019 [11]** | Yes | Yes | NR | Yes | Yes | No | No | Yes | Yes | No | Yes | NA | NA | Yes | Good |
| **Ramos et al., 2017 [12]** | Yes | Yes | NR | Yes | No | No | No | Yes | Yes | No | Yes | NA | NA | No | Fair |
| **Chung et al., 2018 [13]** | Yes | Yes | No | Yes | No | No | No | No | Yes | No | Yes | NA | NA | Yes | Fair |
| **Idler et al., 2015 [14]** | Yes | Yes | Yes | Yes | No | No | No | No | Yes | No | Yes | NA | NA | No | Fair |
| **Solomon-Moore, 2019 [15]** | Yes | Yes | Yes | Yes | No | No | No | No | Yes | No | Yes | NA | NA | No | Fair |
| **Bringolf-Isler et al., 2010 [16]** | Yes | Yes | Yes | Yes | No | No | No | No | Yes | No | Yes | NA | NA | No | Fair |
| **Lévesque et al., 2016 [17]** | Yes | Yes | Yes | Yes | Yes | No | No | Yes | Yes | No | Yes | NA | NA | No | Good |
| **McMillan et al., 2016 [18]** | Yes | Yes | Yes | Yes | No | No | No | No | Yes | No | Yes | NA | NA | Yes | Good |

| | | | | | | | | | | | | | | |
|---|---|---|---|---|---|---|---|---|---|---|---|---|---|---|
| **Fan et al., 2019 [19]** | Yes | Yes | Yes | Yes | No | No | No | Yes | Yes | No | Yes | NA | NA | No | Good |
| **da Costa et al., 2020 [20]** | Yes | Yes | Yes | Yes | No | No | No | Yes | Yes | No | Yes | NA | NA | No | Good |
| **Park and Lee, 2020 [21]** | Yes | Yes | Yes | Yes | Yes | No | No | Yes | Yes | No | Yes | NA | NA | No | Good |
| **Vandermeerschen et al., 2015 [22]** | Yes | Yes | Yes | Yes | No | No | Yes | No | Yes | No | No | NA | NA | No | Fair |
| **Paudel et al., 2014 [23]** | Yes | Yes | Yes | Yes | No | No | No | No | Yes | No | No | NA | NA | Yes | Fair |
| **Wang and Qi, 2016 [24]** | Yes | Yes | Yes | Yes | No | No | No | Yes | Yes | No | Yes | NA | NA | No | Good |
| **Macniven et al., 2016 [25]** | Yes | Yes | No | Yes | No | No | No | No | Yes | No | Yes | NA | NA | No | Poor |
| **Haddad et al., 2019 [26]** | Yes | Yes | NR | Yes | No | No | No | No | Yes | No | Yes | NA | NA | Yes | Fair |
| **Badura et al., 2021 [27]** | Yes | Yes | NR | Yes | No | No | No | Yes | Yes | No | Yes | NA | NA | Yes | Good |
| **da Costa et al., 2021 [28]** | Yes | Yes | No | Yes | No | No | No | Yes | Yes | No | Yes | NA | NA | No | Fair |
| **Júdice et al., 2021 [29]** | Yes | Yes | NR | Yes | No | No | No | No | Yes | No | Yes | NA | NA | No | Poor |
| **Kefeli Çol and Altay, 2021 [30]** | Yes | Yes | NR | NR | Yes | No | No | Yes | Yes | No | Yes | NA | NA | No | Fair |
